# Utilization of a Psychosis Consultation Service: Early Lessons from a Statewide Initiative

**DOI:** 10.1101/2025.04.21.25324324

**Authors:** Sümeyra N. Tayfur, Laura A. Yoviene Sykes, Cenk Tek, Vinod H. Srihari

## Abstract

In February 2024, the Program for Specialized Treatment Early in Psychosis (STEP) in Connecticut launched the STEP Learning Collaborative (STEP-LC), a statewide learning health system. As part of this initiative, STEP-LC introduced a free consultation service to support clinicians, administrators, and healthcare leaders in the continuing care of young people with recent-onset psychosis (ages 16–35). As the only coordinated specialty care (CSC) clinic in the state offering this provider-to-provider service, STEP aims to address clinical and systemic challenges by providing expert guidance on medication management, psychotherapy, family engagement, and program development. Consultations are requested through a brief online form publicly available on the STEP-LC website (link). Experts at STEP aim to respond within one business day to initiate discussions regarding the identified case. The service is designed to be flexible, offering one-on-one meetings, integrating consultations into existing team meetings at the requesters’ agencies, and allowing outside organizations to observe STEP operations. To date, 26 consultations have been completed: 24 within the state and 2 from out-of-state. Within-state requests primarily involved direct clinical issues, such as treatment planning, transitions of care, and family support strategies. Out-of-state consultations focused on broader systemic needs, including the development of new early psychosis programs and collaborative networks. Feedback from consultees highlights the value of the service in enhancing the quality and accessibility of psychosis care. This initiative underscores the importance of leveraging specialized expertise to support both individual and systemic efforts in early psychosis intervention.

## Introduction

First-episode psychosis (FEP) requires timely and specialized interventions to prevent long-term disability and improve outcomes (e.g., Bird et al., 2018). While early intervention programs have shown significant benefits, many clinicians and healthcare leaders lack access to specialized resources (O’Connell et al., 2021; Moe et al., 2018). A key systemic barrier to help-seeking in FEP is the variability in provider expertise, which can delay appropriate treatment (Causier et al., 2024; Moe et al., 2018). Workforce development is a critical challenge in implementing coordinated specialty care (CSC), involving the recruitment, retention, training, and support of interdisciplinary teams to ensure timely symptom identification and treatment (Powell et al., 2020). High turnover, job-related stress, and training gaps remain as persistent barriers, emphasizing the importance of strategic recruitment, comprehensive training, and strong leadership support (Powell et al., 2020). Consultation services offer a scalable solution by offering targeted support to clinicians, ensuring they receive the guidance and resources necessary for effective early psychosis care.

In February 2024, the Specialized Treatment Early in Psychosis (STEP) clinic in Connecticut launched the STEP Learning Collaborative (STEP-LC), a statewide learning health system designed to improve an explicit set of population health outcomes for young adults (ages 16-35) with recent-onset schizophrenia-spectrum disorders (Srihari et al., 2016). STEP-LC focuses on both improving *access* (measured by the Duration of Untreated Psychosis (DUP) and the quality of regional pathways to care) and care *quality* (measured by a range of patient-centered outcomes). The learning health system operates through distinct workstreams: workforce development, early detection, informatics, stakeholder engagement, sustainability, and care model refinement. The rationale, design, and implementation of this statewide system is described elsewhere (Sykes et al., 2025).

The workforce development workstream built on a long history of training initiatives by the STEP clinic, provided to various stakeholder groups through both ad hoc invitations and proactive outreach. These efforts also supported a previous NIH-funded DUP reduction campaign from 2015 to 2019 (Srihari et al., 2014). The cumulative experience of this work enabled the rapid launch of STEP-LC’s workforce development efforts, offering tailored education for providers and community members. Provider trainings included a six-session course on early intervention for schizophrenia (link) and monthly sessions (link) that evolved from ECHO-based (Extension for Community Healthcare Outcomes) case discussions to expert-led presentations on clinician-requested topics (Arora et al., 2011). Family and community workshops (link) provided education tailored to caregivers while also engaging the broader community as potential supporters in a patient’s care journey. Resources, including slides and webinar recordings, were posted on the STEP-LC website for asynchronous learning, and attendees were encouraged to share them in the public domain, with many available on YouTube (MindmapCT and resource library).

Based on provider needs and the STEP team’s experience, a consultation service was launched to support clinicians, administrators, and healthcare leaders in caring for young people with FEP (ages 16–35). The demand for informal, ad hoc access to experienced peers at STEP emerged through workforce development activities and STEP-LC staff processing referrals. Clinicians attending webinars or ECHO sessions found scheduled events challenging and needed more detailed case discussions. Similarly, providers making referrals—especially for ineligible patients—valued informal consultations on access and care management challenges.

The STEP Consultation Service was created as a provider-to-provider channel, specifically designed to lower barriers for community clinicians to access STEP’s multidisciplinary team. Operating alongside professional training initiatives and educational workshops, it reinforces a commitment to continuous learning and quality improvement. As the only CSC clinic in the state offering this provider-to-provider service, this paper outlines the establishment and early utilization of the STEP Consultation Service, highlighting the challenges addressed and key insights gained during its first year of implementation.

## Method

The STEP Consultation Service provides guidance on challenges across all levels of care, including frontline clinician concerns (medication management, psychotherapy, family engagement, and education or employment support), opportunities for managers to improve healthcare processes and quality, and strategies for healthcare leaders seeking to enhance or develop services for young adults with recent-onset schizophrenia. A brief online form, specifically developed for this service, is publicly available on the STEP-LC website for consultation requests (link). Experts at STEP aim to respond within one business day to initiate discussions on the identified case. The service is designed for flexibility, offering one-on-one meetings, integrating consultations into existing team meetings at requesters’ agencies, and allowing outside organizations to observe STEP operations. This project is part of the STEP-LC service implementation, which is approved by the Yale University Institutional Review Board under the Yale Human Research Protection Program (New Haven, CT, USA). As this is not a separate research study, no additional ethical approval was required. Details of the approved implementation protocol are available in Sykes et al. (2025). No protected health information (PHI) is shared during consultations.

### Service Design and Scope

The consultation service supports clinicians, administrators, and healthcare leaders in the continuing care of individuals aged 16–35 experiencing FEP. The service includes a structured consultation request form that excludes PHI about individuals under care. Reason for consultation (medication, psychotherapy, family services, education or employment support, building a new service), consultee type (clinician or administrator), how they heard about the service, age range of the patient (<18 or 18-35), town where the patient resides, and contact information of the consultee are captured in the request form. Alerts about requests submitted through the website are sent to a shared email account, allowing designated STEP staff to distribute and track consultation requests. Individuals seeking treatment or making referrals on behalf of individuals with psychosis are redirected to the centralized STEP-LC referral line. Consultations from clinical staff working with Connecticut residents are prioritized for rapid response, while requests from outside the state or country are directed to a separate email inbox and reviewed weekly.

### Data Collection and Reporting Outcomes

All consultation requests are logged in an Excel spreadsheet, which captures the information from the request forms including the date of request, reason for consultation, type of consultee, referral source, patient age range (<18 or 18–35), town of residence, and consultee contact information as well as outcomes and case notes from the consultation. Descriptive statistics are used to summarize consultation requests and outcomes. Consultation notes and feedback from consultees are used to highlight key themes, common challenges, and gauge the service’s impact.

## Results

Since its launch, the STEP Consultation Service has received a total of 26 consultation requests via the online portal, with 24 (92.31%) occurring within Connecticut and 2 (7.69%) from out-of-state. The in-state consultations were case consultations, while the out-of-state requests were administrative in nature. Of the 24 case consultations, 10 (41.67%) involved minors (patients aged 16-18). Requests came from across the state, with the highest number (n=6; 25%) originating from the southwest region of Connecticut. Most referrals within the state came from clinicians who heard about the service through another provider, clinician, or agency (n=13; 54.17%). Of the 26 requests, 21 (80.77%) consultations were successfully completed, with several leading to ongoing support or follow-up. Feedback from consultees was positive, highlighting the service’s value in addressing complex clinical cases and improving care coordination. Despite clear messaging on the service’s focus on FEP, some consultations involved patients with chronic schizophrenia. Additionally, consultations remained incomplete (n=5; 19.23%) for the same reason—requesters did not respond to repeated outreach via phone and email.

The majority of the case consultations were focused on direct clinical care, including medication management, psychotherapy or family engagement, and reflected a need for guidance in complex cases that often cut across these categories. In contrast, the out-of-state consultations were administrative in nature, primarily requesting guidance about developing early psychosis services and establishing learning collaboratives.

Despite overlapping reasons, **psychotherapy and family support** represented the most common consultation focus (Table 1). Many inquiries sought strategies for engaging families in treatment, improving patient adherence to therapy, and structuring effective psychotherapeutic interventions. Family engagement was a recurring theme, as many clinicians encountered challenges in aligning treatment goals between patients and their caregivers, particularly in cases where families were hesitant about psychiatric interventions.

**Table 1.** Reasons for Requesting Consultation *Note*. Some requests were made for multiple reasons, so the total does not sum to 26 or 100%. Those who selected “Other” as their reason often described issues that overlapped with existing categories.

| **Reason** | **n (%)** | **Examples** |
| --- | --- | --- |
| Medication management | 10 (38.46%) | Antipsychotic selection, dosing, treatment resistance |
| Psychotherapy | 14 (53.85%) | Engaging families, therapy structuring, adherence issues |
| Family services | 8 (30.77%) | Discharge planning, referral guidance, youth-to-adult transitions |
| Program development | 2 (7.69%) | Creating early psychosis programs, cross-agency collaboration |
| Other | 6 (23.08%) | Initial evaluation guidance, diagnostic uncertainty, co-occurring autism-spectrum considerations, addressing legal or administrative barriers, working with clients in residential settings, developing tools for crisis |

**Medication management and optimization** were the second most common consultation requests. These cases typically involved guidance on selecting appropriate antipsychotic treatments, optimizing medication dosages, and managing treatment-resistant psychosis. Given the complexity of psychotropic medication regimens, clinicians frequently sought expert input on balancing efficacy with potential side effects, particularly for young individuals. Many consultations also surfaced diagnostic uncertainty and the need for more attention to differential diagnosis.

Some consultations focused on **transition planning and disposition**, particularly discharge planning from inpatient settings, post-hospitalization care recommendations, and transitions from youth to adult mental health services. Many of these cases required coordinated efforts across multiple healthcare providers to ensure continuity of care and reduce the risk of treatment disengagement.

Lastly, **program development and healthcare system related issues** were the least common consultation topics, and requested exclusively by out-of-state administrators. These consultations focused on building new early psychosis intervention programs and enhancing care coordination across different healthcare agencies. The interest in system-level consultation underscores the need for structured learning collaboratives and specialized training to support the expansion of evidence-based psychosis treatment models.

### Case Examples

To illustrate the impact of the consultation service, a few case examples are highlighted (key identifiers have been suppressed):

#### Case 1: Medication Management and Family Engagement

*An adolescent female with schizoaffective disorder was referred for consultation following her second psychiatric hospitalization. The referring clinician sought guidance on disposition planning, medication adjustments, and family engagement strategies. The consultation team recommended a structured outpatient treatment plan with an emphasis on psychoeducation for the family. The case highlighted the challenges of coordinating inpatient-to-outpatient transitions and the need for tailored family interventions to enhance adherence and long-term outcomes*.

#### Case 2: Treatment-Refractory Schizophrenia

*A young adult male with a history of psychosis was referred for consultation regarding treatment resistance. The primary challenge involved optimizing antipsychotic management while considering adjunctive psychosocial interventions. The consultation service advised a reassessment of medication response, potential augmentation strategies, and structured cognitive-behavioral therapy for psychosis (CBTp). This case underscored the importance of individualized pharmacologic and psychotherapeutic approaches in managing treatment-resistant psychosis*.

#### Case 3: Engaging Families in Suicidality Risk Management

*A young adult male with schizoaffective disorder and a recent suicide attempt was referred for consultation with concerns about family engagement in safety planning. The consultation team emphasized the need for structured family psychoeducation and crisis planning while addressing the family’s hesitancy. Follow-up consultations facilitated a collaborative approach involving both the individual and their support system, improving engagement and safety monitoring. This case emphasized the value of involving families in risk management strategies*.

#### Case 4: Developing a New Early Psychosis Service

*An administrator from an out-of-state psychosis program sought consultation on building a new coordinated specialty care (CSC) program. The team provided guidance on evidence-based models, staffing considerations, and funding strategies. Additionally, discussions explored incorporating digital mental health tools and community partnerships to enhance service accessibility. This case highlighted the expanding role of consultation services in supporting systemic improvements in early psychosis care*.

## Discussion

The early experiences of our consultation service have provided key insights into the needs of clinicians and administrators seeking expert guidance in psychosis care. Consultations primarily focused on direct patient care, with many consultees requesting support in medication management, psychotherapy, and family services, highlighting the need for specialized expertise. The positive feedback from consultees suggests that the consultation service effectively meets a clinical need, particularly for complex cases requiring specialized guidance. Despite the service’s focus on FEP, the inclusion of chronic schizophrenia cases suggests a need for broader clinical guidance, particularly in managing long-term, treatment-resistant cases. Additionally, incomplete consultations may indicate a need for a more streamlined referral process or alternative communication methods to improve accessibility and follow-through.

Many of the requests helped identify family engagement as a key challenge. Clinicians frequently reported difficulties in involving caregivers who were reluctant or lacked understanding of psychotic illness or had developed a strained relationship with their child or with the treatment team. Many also felt limited by their own lack of experience and time to adequately assess and address these issues. Given the well-established role of family psychoeducation in improving treatment adherence (Iuso et al., 2023), these findings underscore the need for innovative ways to ease delivery of family interventions while expanding the capabilities of clinical providers and agencies. As part of its efforts, STEP-LC will work to facilitate family psychoeducation by offering statewide virtual sessions that provide culturally responsive support tailored to diverse family needs. Early, proactive outreach and clear communication about the benefits of family involvement could further enhance participation and improve outcomes for individuals with FEP. A recent systematic review found that, compared to usual care, family interventions significantly reduced carer burden, psychological distress, and expressed emotion, while also moderately decreasing patient hospitalization rates, underscoring their benefits for both caregivers and patients (Gleeson et al., 2025).

The two out-of-state inquiries around service development are consistent with an increasing number of requests to our program and reflect a growing recognition of the promise of early intervention services for schizophrenia spectrum disorders. The STEP clinic has consulted and collaborated with several innovative efforts to implement learning health systems outside Connecticut, (Breitborde et al., 2022; Vohs et al., this issue; Mathis et al., this issue). This has resulted in a virtuous cycle between lessons learned within the STEP-LC and across diverse state mental healthcare systems.

A growing number of online resources provide valuable information on early intervention (EPINET, n.d.), but we are unaware of a service like the one described here. The STEP Consultation Service offers a low-barrier, provider-to-provider approach, removing the need for formal case narratives or predefined questions. Instead, it fosters dynamic discussions, helping experts refine and expand the questions that prompted the initial request. This model ensures immediate, flexible guidance to enhance provider decision-making, while also informing workforce development topics and strengthening goodwill among referral sources—both critical for advancing statewide efforts to improve access and care quality.

## Future Directions

### Modifications to improve accessibility of the service

To enhance accessibility and streamline the consultation process, we implemented a more efficient request system to address the recurring issue that resulted in five incomplete consultations. Previously, scheduling a consultation often required multiple exchanges between consultees and experts, leading to delays and increased administrative burden. To reduce these inefficiencies and eliminate the need for repeated attempts to reach the appropriate contact, we transitioned to Microsoft Bookings (Microsoft, n.d.) as our primary scheduling platform. This new system allows consultees to directly select and book a virtual or phone appointment with one of our staff members at their convenience. We also provide brief introductions to our staff and their areas of expertise, enabling requesters to select the most relevant expert. By automating the scheduling process, we aim to improve response times, increase service accessibility, and reduce logistical challenges, ultimately enhancing the overall efficiency of the consultation service.

### Promotion of the service through professional outreach and awareness initiatives

The early detection activities planned as part of STEP-LC will more deliberately promote the STEP consultation service, which will remain available without cost to the healthcare providers and systematically document all consultative interactions. The STEP-LC will promote the consultation service as part of an ongoing statewide Early Detection campaign (link), which aims to reduce the duration of untreated psychosis (DUP) and improve pathways to care (Sykes et al., 2025). This component of STEP-LC employs a three-pronged approach. The first focuses on public awareness through mass media and digital outreach, including television news appearances, press releases, transit advertisements, billboards, radio ads, and targeted social media campaigns. Search engine optimization strategies enhance the visibility of the STEP-LC website, which includes the consultation service portal. The second approach engages clinical and non-clinical stakeholders through professional outreach and detailing. Activities include educational presentations, individual meetings with referral sources, and distribution of branded materials such as brochures, stress balls, pens, stickers, and water bottles (e.g., brochures that promote the consultation service). The third approach ensures rapid access to services through STEP-LC’s centralized referral line, staffed by Early Detection and Assessment Coordinators (EDACs), who screen individuals and facilitate expedited referrals. In addition to formal referrals, EDAC interactions often involve ad hoc consultative discussions that address immediate clinical concerns. These real-time consultations meet the urgent needs of providers or families, potentially reducing the number of formal consultation requests while still enhancing access to expert guidance. The relatively modest volume of requests received via the online referral form are thus an underestimate of the true number of consultative interactions, and increased efforts will be made to document these going forward to allow for a more accurate evaluation of impact.

### Integrating the consultation service with other workflows in STEP-LC

While the consult service fits naturally within the menu of workforce development activities described earlier, there are important links to other activities that we expect to deepen as the learning health system matures. For example, a key goal of the informatics platform (Mathis et al., this issue) is to leverage population outcomes for quality improvement across the statewide network. Implementing data-driven changes in care will require training and support for clinicians and clinical managers. While webinars and formal sessions will help, the consultation service is expected to become an important portal to serve as both a reinforcement tool and a more flexible, individualized avenue for education and support. Similarly, consultations for cases that do not respond to available care will raise the need for refinements in care models (e.g., adding a supported employment specialist to a team), research to address gaps in current treatment models (e.g., addressing negative symptoms), or adaptation of interventions for delivery in community settings (e.g., online cognitive remediation).

### Evaluation of the consultation service’s performance and impact

We will continue tracking the consultation service’s performance to inform its potential scalability and sustainability, ensuring it remains a valuable resource for advancing psychosis care at both individual and organizational levels. As part of a broader intervention measuring population health outcomes, there will be an opportunity to explore its specific contribution, particularly in enhancing provider competence in early psychosis care. Experts in continuing education have long emphasized workplace-based learning as essential for applying knowledge acquired through traditional teaching formats (Wyatt, 2005). Learning that happens in real time, addressing clinicians’ immediate uncertainties, is often more effective than scheduled classroom or webinar-based instruction but remains underutilized. By embedding the consultation service within a learning health system, we aim to overcome barriers to ad hoc learning and support clinicians in developing professional judgment (Coles, 2002). Initial feedback suggests improvement in provider comfort in caring for psychosis patients after interaction with our staff. Further work will help define measures that will allow us to continuously improve and evaluate impact on professional development.

## Conclusion

This review of the first year of a provider focused consult service confirms proof of concept and provides optimism to expand utilization and develop measures to more fully evaluate impact in the context of a new statewide learning health system for recent onset schizophrenia. Queries came from healthcare professionals across the state and in a variety of roles. The questions confirmed interest and need for consultation on key aspects of early intervention service models and users informally reported a high degree of satisfaction and increased confidence in the care of psychosis patients. The range of requests underscore the critical role of expert consultation in supporting clinicians, administrators, and healthcare leaders in optimizing both direct clinical care and service design. Notably, the STEP team responded to clinical questions across the illness course, suggesting educational needs even amongst providers caring for longstanding patients with psychotic disorders. Continued evaluation and refinement of the service will inform its future development and broader implementation within a statewide learning health system for first-episode psychosis.

## Data Availability

All data produced in the present work are contained in the manuscript

## Acknowledgements

We thank the clinicians and administrators who utilized the STEP Consultation Service and provided valuable feedback.

